# COVID-19 in Uganda: Predicting the impact of the disease and public health response on disease burden

**DOI:** 10.1101/2020.05.14.20102202

**Authors:** David Bell, Kristian Schultz Hansen, Agnes N. Kiragga, Andrew Kambugu, John Kissa, Anthony K. Mbonye

## Abstract

**Objective:** COVID-19 transmission and the public health ‘lock-down’ response are now established in sub-Saharan Africa, including Uganda. Population structure and prior morbidities differ markedly between these countries from those where outbreaks were previously established. We predicted the relative impact of COVID-19 and the response in Uganda to understand whether the benefits could be outweighed by the costs.

**Design and setting:** Age-based COVID-19 mortality data from China were applied to the population structures of Uganda and countries with previously established outbreaks, comparing theoretical mortality and disability-adjusted life years (DALYs) lost. Based on recent Ugandan data and theoretical scenarios of programme deterioration, we predicted potential additional disease burden for HIV/AIDS, malaria and maternal mortality.

**Main outcome measures:** DALYs lost and mortality.

**Results:** Based on population age structure alone Uganda is predicted to have a relatively low COVID-19 burden compared to equivalent transmission in China and Western countries, with mortality and DALYs lost predicted to be 12% and 19% that of Italy. Scenarios of ‘lockdown’ impact predict HIV/AIDS and malaria equivalent to or higher than that of an extensive COVID-19 outbreak. Emerging HIV/AIDS and maternal mortality data indicate that such deterioration could be occurring.

**Conclusions:** The results predict a relatively low COVID-19 impact on Uganda associated with its young population, with a high risk of negative impact on non-COVID-19 disease burden from a prolonged lockdown response. The results are likely to reflect the situation in other sub-Saharan populations, underlining the importance of tailoring COVID-19 responses to population structure and potential disease vulnerabilities.

**Transparency statement:** The lead author affirms that the manuscript is an honest, accurate, and transparent account of the study being reported; that no important aspects of the study have been omitted; and that there are no discrepancies from the study as originally planned.

## Introduction

The COVID-19 pandemic is redirecting focus and prioritization of health systems globally. Public health responses have been dominated by enforced social distancing and stay-at-home interventions, characterized as ‘lockdowns’ advocated by the World Health Organization (WHO).[1] As reported SARS-CoV-2 infections increase on the African continent, countries are implementing measures including closure of workplaces and severe restrictions on travel, implicated in reducing transmission and pressure on acute healthcare facilities elsewhere.[2] Reducing close contact between people should reduce the basic reproduction rate, R_0_, and overload of health services.[3]

Relatively few cases of COVID-19 have been recorded to date in sub-Saharan African countries.[4] However, sub-Saharan Africa is considered particularly vulnerable to COVID-19 due to relatively weak health service infrastructure and low clinician-population ratios,[5] relatively limited laboratory capacity,[6] and a higher rate of underlying conditions including malnutrition, anemia, HIV/AIDs, and chronic respiratory conditions due to tuberculosis and air pollution.[7] There is paucity of data on interactions between COVID-19 and other acute febrile disease including malaria, which could exacerbate or ameliorate the SARS-CoV-2 infection.[8] These same conditions increase the population’s dependence on health service access and the commodity supply lines that support it, including access to long-term medication and care in acute illness. Access is health services is also essential for reducing poor antenatal and birthing outcomes.

Health services are at risk in the COVID-19 outbreak through various mechanisms. Clinicians and direct care givers of COVID-19 patients have a disproportionately higher mortality than the general age-adjusted population.[9] Closure of logistics-related workplaces and transport services interrupts supply lines.[10] Travel bans and reduction in public transportation limits access, public perceptions of increased risk of SARS-CoV-2 infection near health facilities dissuade attendance, while clinic activities considered ‘non-urgent’, such as ante-natal care, may be closed.[11]

In contradistinction to potential health vulnerabilities, sub-Saharan African countries could be ‘protected’ from COVID-19 mortality by a very different age structure compared to countries where mortality has been particularly high, such as Italy, Spain, the USA and in Hubei Province in China.[12,13] Countries with far higher proportions of children to older adults are expected to have lower overall mortality and higher rates of asymptomatic infection.[14] Age comparisons are of particular importance to public health, as Disability-Adjusted Life Year (DALYs), fundamental to assessing relative burden, are highly influenced by age of death or onset of long-term disability.[15]

Uganda typifies many of the characteristics of sub-Saharan countries. With a fertility rate above 5·5% of the population are estimated to be below 17 years of age and only 1.6% are ≥70 years in 2020.[12,16] Incidence and mortality from malaria (12,356,577, 13,203) and tuberculosis (86,000 and 19,000) respectively in 2018 imparted a significant burden.[17,18] Maternal mortality ratio was 368 per 100,000 live births in 2016,[19] while 100,000 HIV positive women were dependent on antiretroviral therapy (ART) to prevent mother to infant transmission in 2018.[20] Success of the HIV/AIDS program has reduced AIDS-related mortality from 43,000 in 1990 to 23,500 in 2018,[20] resulting in a growing number of people living with HIV (1,400,000 in 2018) and dependent on frequent monitoring, anti-retroviral therapy and treatment of co-infections.[20] With low ratios of physicians to population (0.1/1000 people) and hospital beds (0.5/1000),[21] Uganda has a relatively fragile health system with limited capacity to expand critical care services.

In view of this, we hypothesize that mortality in sub-Saharan Africa accrued from COVID-19 and from interruption of non-COVID-19 disease programs that result from the response will differ greatly from those of outbreak countries in Europe, North America and east Asia. As of May 02, 2020, Uganda had conducted 29,277 tests for COVID-19, and of these 85 (0.29%) tested positive, while 52 persons have recovered, and no deaths have been registered.[22] The neighboring countries of Kenya and Tanzania have community transmission with close to five-hold higher numbers of covid-19 cases,[23] posing a threat of importation.

We therefore predicted the potential burden of COVID-19 in Uganda based on its population age structure in comparison to an equivalent outbreak in other countries with established COVID-19 transmission. We calculated mortality and Disability-Adjusted Life Years (DALYs) lost, a summary measure of life-time impact combining time lost through premature death and time lived in states of less than optimal health.[15] We then predicted the potential for impact of a COVID-19 lockdown response on the burden of selected diseases in Uganda, where restrictions on travel and workplace attendance have been implemented by the Government of Uganda since March 12^th^, 2020. We grounded this against limited publicly-available data on changes in health burden in Uganda over recent months.

## Methods

Data collection was conducted in Uganda, which has a sporadic transmission of COVID-19 and conditions generalizable across several sub-Saharan countries.[4]

### Data sources to assess recent trends in disease burden

Information on the Uganda population structure population was obtained from the Uganda Bureau of Statistics census report, while data on comparison countries was obtained from UN data (Table S1).[13,24]

Data on HIV and Tuberculosis were obtained from the aggregated President’s Emergency Plan for Aids Relief (PEPFAR) weekly surge reports generated from data across 13 PEPFAR Uganda implementing partners,[25] including the persons newly diagnosed with HIV per the Uganda national HIV/AIDS treatment guidelines.[26] Additional data on age-stratified HIV prevalence were obtained from the Uganda Population-based HIV Impact Assessment (UPHIA) final report.[27]

Age-stratified malaria incidence for 2019-2020 was obtained from the Uganda Health Management Information System (HMIS) quarterly reporting.[28] Maternal mortality data including deliveries from January 2019 to March 2020 inclusive were similarly obtained from the Uganda Health Management Information System (HMIS) quarterly reporting.[28]

### Calculation of the number of DALYs lost

Calculations of DALYs lost followed broadly the methods outlined for the most recent WHO global burden of disease estimates.[29] This involved defining life years lost by age as the difference between actual age of death and the life expectancy in a standard life table reflecting the highest life expectancy in the world today and a set of disability weights to reflect the relative severity of diseases. The unequal age-weighting function as well as discounting of future life years applied in earlier DALY versions were excluded.[15]

COVID-19 disease burden and excess HIV, malaria and maternal mortality were calculated by multiplying the DALYs lost for a single health event by the population incidence and deaths by age group. DALYs for COVID-19 are based on age-related mortality reported by Verity et al. from the China outbreak, applied to age structures of comparison countries with relatively high COVID-19 burden (United States, China, Italy and Spain) and Iceland,[30] where testing rates have been relatively high.[23] For sake of comparison, a 20% total detectable infection rate was applied across all age groups, this being assumed to be a worst case for comparison with deterioration in non-COVID disease burden..

The details of assumptions involved in DALY calculations are provided in the supplementary file. Potential impact of reduced health service access in Uganda through the COVID-19 response was predicted for HIV/AIDS, malaria, maternal mortality. HIV/AIDS predictions assumed an arbitrary low (20%) loss from follow-up (no medication) for current infections extending for 6 months, with mortality returning to 1990 levels (essentially pre-antiretroviral therapy). Reduced detection of new HIV infections and initiation of management is based on first quarter 2020 data.[25]

Excess malaria burden was estimated based on 6 months of incidence and mortality rate changes recently predicted by WHO, for three scenarios of minor, moderate and major reduction in services (WHO scenario 1 (WS1): No insecticide-treated net (ITN) campaigns, continuous ITN distributions reduced by 25%, WS4: No ITN campaigns, access to effective antimalarial treatment reduced by 25%, and WS9: No ITN campaigns, both continuous ITN distributions and access to effective antimalarial treatment reduced by 75%).[31] Relative malaria mortality and incidence rates by age for Uganda were derived from the Institute of Health Metrics data,[32] with the 2018 baseline mortality reported by WHO.[18]

Maternal mortality was based on Uganda data from 2019-2020 and include only mortality, not persisting injury (see Results section).

### Ethical considerations

The study used publicly available secondary aggregate level data. No individual person-identifying information was used. No ethical approval was therefore required.

### Patient and public involvement

Ugandan and other data used in this study are publicly available. There was no direct public or patient involvement in the related work.

## Results

### Trends in reported disease data

#### HIV/AIDS

Identification of new HIV cases declined by 75% in the first 2 weeks of April, with an associated 75% reduction in initiation of izoniazid-preventive therapy (IPT) to prevent secondary tuberculosis.[25] These dips were similar to a temporary decline over the 2019-2020 Christmas - New year period that subsequently recovered (Figure 1, Table S2).

**Figure 1.**
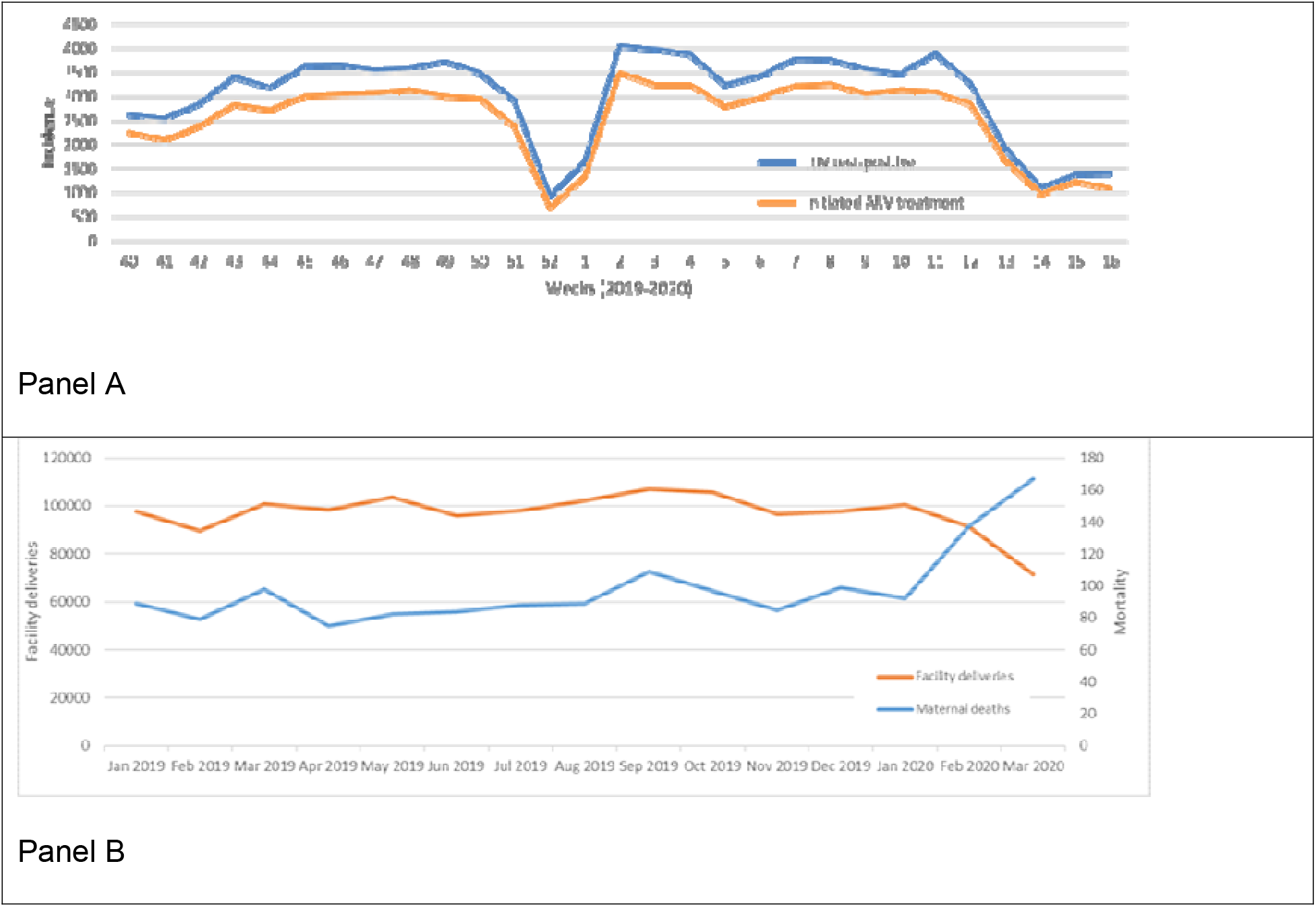
Panel A. HIV incidence and initiation of antiretroviral therapy (ART) in Uganda, week 40 (October 2019) to week 16 (end-April) 2020.[25] Panel B. Facility deliveries and maternal mortality, Uganda, 2019 to end- March 2020.

#### Malaria

Malaria showed a reduction in case detection in the first quarter of 2020, consistent with a seasonal decline but more persistent and of greater magnitude than 2019 (1,206,606, 1,010,524 and 678,176 cases in January, February and March 2020, vs. 832,499, 666,493 and 722,370 in 2019). Admissions and inpatient deaths declined by similar proportions (Table S3, Figure S1).

#### Maternal mortality

A 29% (28,939) reduction in facility deliveries is recorded in Ministry of Health Uganda data in March compared to January 2020, 28% below the 12-month average for 2019. Over the same period, an 82% increase in maternal mortality was recorded (from 92 to 167 women), an increase of 87% over the 12-month 2019 average of 89.5 (Figure 1, Table S4).

### Demography, predictions of DALYs lost and mortality

#### COVID-19

Data in 10-year age groups for Uganda and comparator countries, demonstrate a vastly different population age distribution (Figure 2). Assuming an equal (20%) infection rate in all countries and an age-related mortality rate based on China data,[33] a far lower disease burden is predicted for Uganda than all the comparison countries, with mortality 0.13 and DALYs lost 0.19 that of Italy per head of population (Figure 2, Table 1). The bulk of the COVID-19 burden in Uganda is predicted to fall in late middle-age, peaking in the 60 - 69 year age bracket (Table 2).

**Figure 2.**
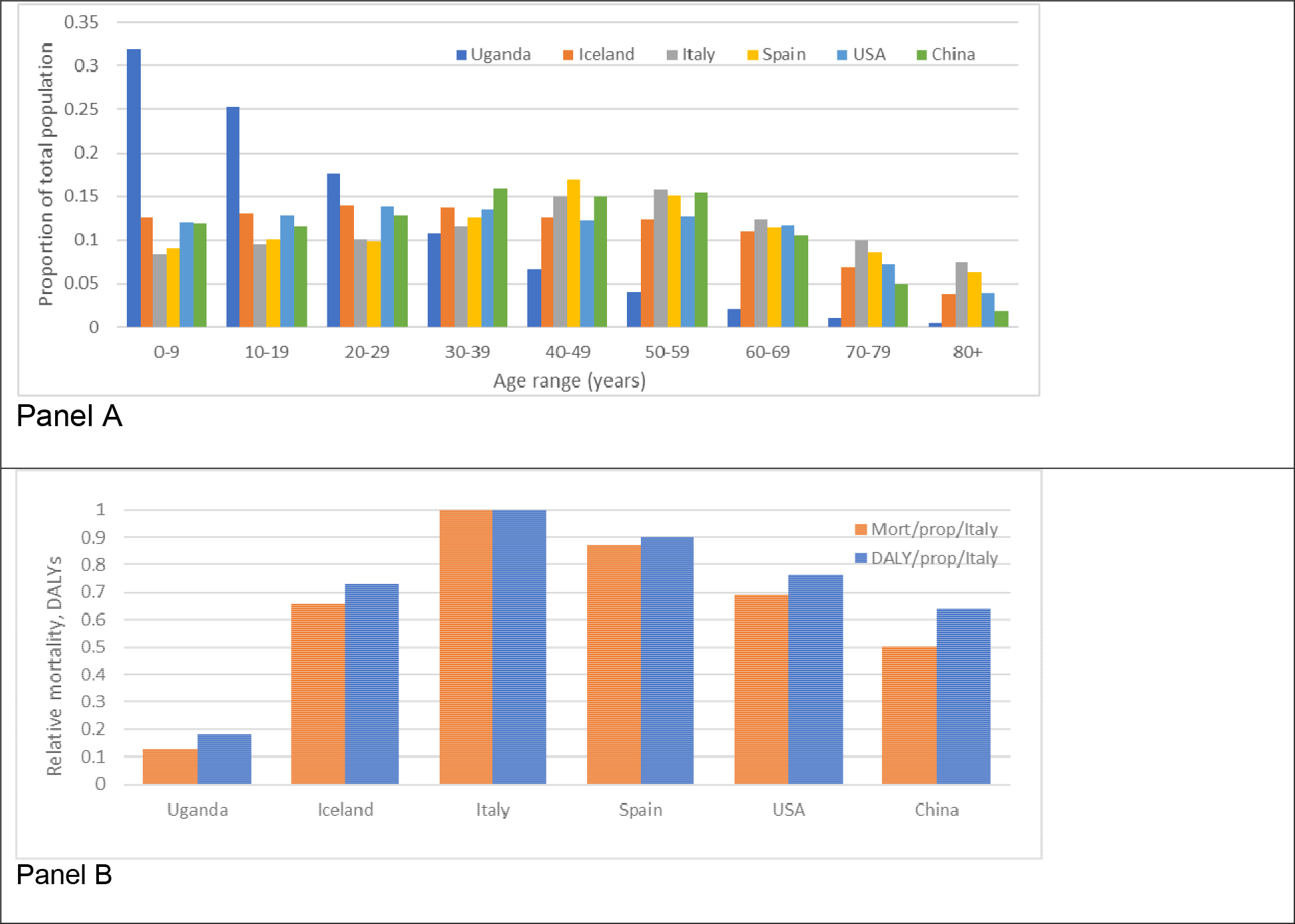
Panel A. Comparison of age profiles of Uganda with countries with COVID-19 outbreaks previously recorded and instituting varying degrees of physical distancing policies: China, USA, Spain, Italy and Iceland. Panel B. Relative mortality and burden of DALYs from COVID-19 predicted by age distribution alone, assuming an equivalent infection rate per country, standardized against the burden predicted for Italy. Age-specific IFR is based on China.[33]

**Table 1.**
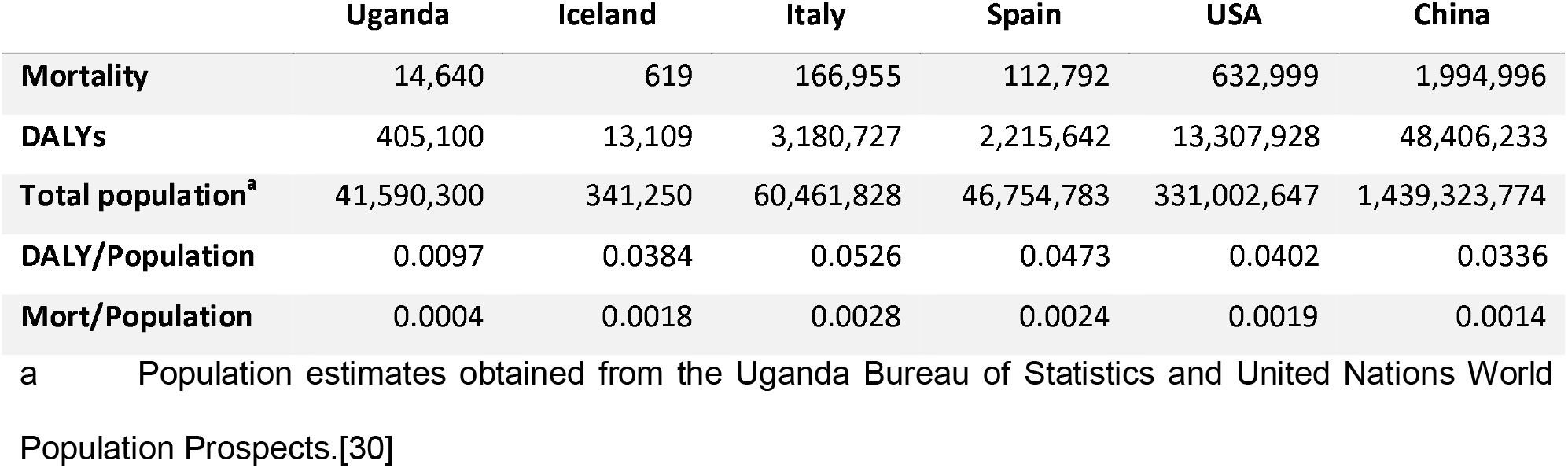
Predicted mortality and DALYs lost for Uganda and comparator countries, based on age-related mortality from the China outbreak, and assuming a 20% detectable infection rate.

|  | Uganda | Iceland | Italy | Spain | USA | China |
| --- | --- | --- | --- | --- | --- | --- |
| **Mortality** | 14,640 | 619 | 166,955 | 112,792 | 632,999 | 1,994,996 |
| **DALYs** | 405,100 | 13,109 | 3,180,727 | 2,215,642 | 13,307,928 | 48,406,233 |
| **Total population<sup>a</sup>** | 41,590,300 | 341,250 | 60,461,828 | 46,754,783 | 331,002,647 | 1,439,323,774 |
| **DALY/Population** | 0.0097 | 0.0384 | 0.0526 | 0.0473 | 0.0402 | 0.0336 |
| **Mort/Population** | 0.0004 | 0.0018 | 0.0028 | 0.0024 | 0.0019 | 0.0014 |

a Population estimates obtained from the Uganda Bureau of Statistics and United Nations World Population Prospects.[30]

**Table 2.** DALYs lost in Uganda assuming a 20% detectable infection rate of SARS-CoV-2.

| Age range (years) | Age of death (years) | Number of deaths<sup>a</sup> | DALYs lost |
| --- | --- | --- | --- |
| 0 - 9 | 5 | 42 | 3,696 |
| 10 - 19 | 15 | 147 | 11,355 |
| 20 - 29 | 25 | 452 | 30,320 |
| 30 - 39 | 35 | 755 | 43,169 |
| 40 - 49 | 45 | 892 | 42,180 |
| 50 - 59 | 55 | 2,028 | 76,028 |
| 60 - 69 | 65 | 3,293 | 91,733 |
| 70 - 79 | 75 | 3,965 | 73,836 |
| 80+ | 85 | 3,064 | 32,783 |
| **Total** |  | 14,640 | 405,100 |

a Based on mortality in China.[33]

#### HIV/AIDS

Assuming a 20% loss to follow-up for 6 months among people living with HIV and a consequent return to 1990 mortality levels for this population, and assuming an equal mortality across age groups, 463,168 excess DALYs are predicted to be lost (Table S5A). The burden from undiagnosed new HIV cases, based on the 55% decline in new ARV treatment initiation in April 2020 and using standard assigned HIV DALYs, predicts 41,757 excess cases failing to commence therapy over 6 months, with 12,151 DALYs lost (Table S5B). Combined, this predicts 475,319 DALYs lost (Table 3)

**Table 3.** Comparison of predicted DALYs lost from COVID-19 if national detectable infection rate reaches 20%^a^, and potential response vulnerabilities over 6 months, based on assumptions as listed.

| Cause | Basis of prediction | Excess DALYs lost | Notes |
| --- | --- | --- | --- |
| **COVID-19** | 20% incidence | 405,100 | 20% total detectable incidence nationally |
| **HIV/AIDS** | Missed new diagnoses<sup>b</sup> and loss to follow-up | 475,319 | Standard HIV DALYs. Missed new treatment initiation based on March 2020 vs. 2019 6-month average and assumed 20% loss to follow-up reverting to 1990 HIV mortality rate. |
| **Malaria** | WHO scenario 1<sup>c</sup> | 257,780 | No ITN campaigns, continuous ITN distributions reduced by 25% |
|  | WHO scenario 4<sup>c</sup> | 509,393 | No ITN campaigns, access to effective antimalarial treatment reduced by 25% |
|  | WHO scenario 9<sup>c</sup> | 2,427,769 | No ITN campaigns, both continuous ITN distributions and access to effective antimalarial treatment reduced by 75% |
| **Maternal mortality** | March 2020 mortality rate<sup>b</sup> | 31,343 | March 2020 rate, vs. 2019 6-month average. |

a Mortality for detectable infection rate based on incidence recorded in China,[33] and including only diagnosed and recorded infections.
b Uganda 2020 data. See Results.
c WHO.[18,31]

#### Malaria

The Ugandan population is predicted to be significantly impacted if the WHO scenarios are realized, reflecting the low age at which most mortality occurs. Mortality and total DALYs lost for WHO scenarios being 3,209 and 257,780 (WS1), 6,533 and 509,394 (WS4) and 31,046 and 2,427,769 (WS9) respectively (Table 3 and Table S6A, B).

#### Maternal mortality

Based on the observed increase in maternal mortality in early March 2020 compared to the preceding 2019-2020 average, an excess 486 deaths are predicted for a 6-month period, incurring 31,343 DALYs lost (Table S7). Non-fatal morbidity has not been taken into account.

Relative costs in DALYs lost and mortality for the modelled scenarios are found summarized in Table 3.

## Discussion

Based on population age structure alone Uganda appears relatively protected from severe COVID-19, with an equivalent sized outbreak imparting a far lower predicted COVID-19 burden per head of population than all comparator countries, less than a third that of China to less than a fifth that of Italy. Within Uganda, a very widespread COVID-19 outbreak (20% of population testing positive) is predicted to impart a lower burden than potential lock-down impacts in HIV/AIDs and malaria programmes alone. The fragility of the Uganda HIV/AIDS burden in particular is reflected in the high predicted DALYs lost with moderate reductions in health service access. The assumption of a 20% reduction in HIV/AIDs follow-up is lower than the reduction in new case detection actually demonstrated in the PEPFAR data.

Early data showing rising maternal mortality and reduced facility-based deliveries, and reduced case-finding for HIV/AIDS indicate that deterioration in essential health service access associated with the COVID-19 pandemic is occurring, risking reversal of hard-won gains in addressing fundamental vulnerabilities in women’s and children’s health. As official social distancing policies were only instituted in Uganda since March 12^th^, 2020, the time of impact of the mandatory lockdown policy is limited, though prior knowledge of COVID-19 may have influenced prior health-seeking behavior. Such reduced health service access may explain the prolonged seasonal decline in malaria case finding in 2020, but a longer reporting period needs to be assessed (Figure S1).

This paper has taken a conservative approach to estimates of program deterioration, while assuming high transmission of COVID-19 with a 20% detectable infection rate across Uganda. As ‘detectable infection’ here is based on early China data when testing was scaling up, and subsequent data suggests that asymptomatic SARS-CoV-2 infection is common,[34] the COVID-19 burden predicted here would reflect a population infection rate considerably higher than 20%. It is noted that countries with major established outbreaks are far lower levels of detectable infection overall.[23] The predictions for COVID-19 disease burden used here in comparison to other disease burdens appear very much a worst-case scenario, and probably greatly over-estimate COVID-19 impact.

Several assumptions underlay the predictions for the impact of COVID-19 and the public health response. A significant assumption is that age-related mortality in Uganda would be similar to that of China. The effects of undernutrition, anemia, intestinal parasite infestation, and infectious disease such as TB and HIV are unknown. They are widely assumed to increase susceptibility, as may chronic lung conditions from indoor and outdoor air pollution. However, major risk factors recorded for COVID-19, obesity, diabetes, hypertension and other cardiovascular diseases,[35,36] are generally less prevalent in sub-Saharan populations.[37] In Uganda a recent national survey reported the prevalence of diabetes at 1.4%.[37] It is therefore uncertain whether the population at a given age will be more or less susceptible. Lower access to intensive care, and particularly to oxygen, may reduce recovery rates for severe cases in Uganda and more broadly in this region in DALYs lost and mortality.[38,39] In calculation of DALYs lost from COVID-19, pre-existing morbidities were not taken into account. As those dying from COVID-19 would have had shortened life expectancies due to these conditions, the calculation here probably over-estimates the COVID-19-related burden.

While comparisons with predicted COVID-19 burden for other countries demonstrate a far lower burden expected for Uganda, actual transmission will clearly vary due to factors affecting transmissibility, including population density, social interactions and possible age-related infectivity. These will vary within, as well as between, the countries listed.

The use of DALYs lost, rather than reliance solely on incidence and mortality, seems essential to understand COVID-19 burden. With the burden of COVID-19 heavily concentrated in older age groups,[33,36] each death imparts a far lower burden than is commonly accrued from HIV/AIDs, malaria, or a maternal death. Maternal and HIV deaths not only remove many years of a parent’s life but frequently leave children orphaned, with further resultant costs. The predictions in this paper, though based on fairly modest assumptions of program deterioration from only three disease areas, indicate a considerable risk of collateral damage to other disease programmes compared to the COVID-19 burden from an extensive outbreak predicted here (Table 3). Increases in infant malnutrition and stunting through economic impact of the response, not modelled here, will have impact for years to come. The World Food Program predicts up to 130 million additional people will face acute food insecurity globally, a large proportion in sub-Saharan Africa.[40] The impact of interruption of vaccination programs will increase the longer health service interruptions persist, with outbreaks including measles in neighboring Democratic Republic of Congo a growing threat.[41] An expected 2.1% - 5.1% reduction in GDP across sub-Saharan Africa will impede the ability of health systems and populations to recover,[42] while reduction in support from donor countries is a risk as they deal with economic downturn at home.

Understanding the extent of spread in the community through better testing will be important to understand whether the relatively low COVID-19 numbers recorded in African countries to date are due to late introduction, are an artefact of low testing, or are due to the low rate of severe presentations predicted here. The consequences of getting the equation wrong are dire. Ramping up testing capacity, prioritizing the building of an evidence base on COVID-19 epidemiology in African populations, is urgent if the risks raised in this study are to be minimized.

## Conclusion

The early data from Uganda can be seen as sentinels of reduced health service access. Declines in quality of HIV, malaria and maternal care risk losing years of progress not only through short-term mortality, but through increase in underlying transmission. In this context, the age-based prediction of low COVID-19 impact in Uganda is of particular significance. The predictions presented in this paper strongly suggest that, based on DALYs lost, the impact of the COVID-19 response on single diseases could outweigh the direct impact of an extensive COVID-19 outbreak. Transferring public health responses designed for other populations may not be appropriate without considerable tailoring to local conditions.

A broader modelling approach to COVID-19 is needed, incorporating accepted measures of disease burden rather than solely mortality, and including scenarios for collateral damage in other disease management. Sentinels for deterioration such as facility-based delivery, maternal mortality and case-finding for HIV and tuberculosis should inform interpretation of this modelling. Uganda’s relatively strong surveillance and monitoring networks provide a strong basis for this across a range of diseases.

## Data Availability

All data on which the work is based are publicly available (referenced) while calculations are available from the authors

## Availability of data

All data is in the public domain, and calculations are freely available from the authors

## Author contributions

DB conceived the study, ANK, AK and AKM contributed to the study design and conceptualization, JK and AKM sourced the data, KSH, performed the economic analyses, all authors participated in the drafting, revising and approval of the manuscript.

## Manuscript license

The Corresponding Author has the right to grant on behalf of all authors and does grant on behalf of all authors, a worldwide licence (http://www.bmj.com/sites/default/files/BMJ%20Author%20Licence%20March%202013.doc) to the Publishers and its licensees in perpetuity, in all forms, formats and media (whether known now or created in the future), to i) publish, reproduce, distribute, display and store the Contribution, ii) translate the Contribution into other languages, create adaptations, reprints, include within collections and create summaries, extracts and/or, abstracts of the Contribution and convert or allow conversion into any format including without limitation audio, iii) create any other derivative work(s) based in whole or part on the on the Contribution, iv) to exploit all subsidiary rights to exploit all subsidiary rights that currently exist or as may exist in the future in the Contribution, v) the inclusion of electronic links from the Contribution to third party material where-ever it may be located; and, vi) licence any third party to do any or all of the above.

## Declaration

The lead author (DB) affirms that the manuscript is an honest, accurate, and transparent account of the study being reported; that no important aspects of the study have been omitted; and that any discrepancies from the study as planned (and, if relevant, registered) have been explained.

## Patient and public involvement

There was no direct patient or public involvement in this study. All data used was in the public domain.

List of abbreviations
AIDS: Accquired Immune Defiiency Syndrome
ART: Antiretroviral Therapy
DALYS: Disability-adjusted life years
GDP: Gross Domestic Product
HIV: Human Immunodeficiency Virus
HMIS: Health Management Information System
IPT: Izoniazid Preventive Therapy
ITN: Insecticide-Treated Net
PEPFAR: President’s Emergency Plan for AIDS Relief
SARS-CoV-2: Severe Acute Respiratory Syndrome
UPHIA: Uganda Population HIV Impact Assessment
USA: United States of America
WHO: World Health Organization
WS1: WHO Scenario 1
WS4: WHO Scenario 4
WS9: WHO Scenario 9

